# Multimorbidity Patterns of Chronic Diseases Among Adults in Rural North China

**DOI:** 10.1101/2024.10.18.24315737

**Authors:** Shuai Tang, Yanxing Li, Meili Niu, Zijing Qi, Tianyou Hao, Hongmei Yang, Maoyi Tian, Xinyi Zhang, Xiangxian Feng, Zhifang Li

**Affiliations:** School of Public Health, Shanxi Medical University, Taiyuan 030001, China; Department of Prevention and Health Care, Affiliated Heping hospital of Changzhi medical college, Changzhi, 046000, China; Department of Public Health and Prevention, Changzhi Medical College, Changzhi 046000, China; The George Institute for Global Health, University of New South Wales, Australia; School of Public Health, Harbin Medical University, Harbin 150081, China

**Keywords:** China, rural, adults, multimorbidity

## Abstract

**Background:** The incidence of chronic diseases is increasing, especially in rural areas, where younger patients often exhibit multimorbidity. Understanding multimorbidity in rural adults can guide the development of targeted management strategies for chronic diseases.

**Methods:** This cross-sectional study was conducted in rural North China using whole cluster stratified random sampling to select two counties in Shanxi Province. A total of 2,208 participants aged 30 years or older from 80 villages were enrolled, stratified by gender and age. Data collection involved questionnaires on socio-demographic characteristics, lifestyle, and disease history, along with physical measurements such as height, weight, and waist circumference.

**Results:** Among the 2,208 participants, 58.11% were aged 30-59 years, and 52.17% were female. The prevalence of chronic diseases was 66.53%, with a multimorbidity rate of 32.47%. The most common conditions were hypertension (43.21%), chronic digestive diseases (11.82%), and stroke (10.19%). Multimorbidity was more prevalent in those aged ≥60 years compared to the 30-59 age group (47.68% vs. 21.51%, P<0.05), with no significant gender differences. Hypertension was present in 78.52% of disease patterns, with common dyads being hypertension & stroke (7.47%), hypertension & heart disease (6.25%), and hypertension & diabetes mellitus (6.11%). In the 30-59 age group, hypertension & chronic digestive disease (3.82%) were most prevalent, while for those aged ≥60 years and males, hypertension & stroke were most common (12.65% and 9.47%, respectively). Among females, the most frequent dyad was hypertension & arthritis (8.16%).

**Conclusion:** Chronic diseases and multimorbidity are prevalent in rural North China, primarily driven by hypertension. Multimorbidity patterns differ by age and gender, indicating the need for targeted prevention and treatment strategies.

## Background

With the aging of the population, the prevalence of chronic diseases continues to rise, and the coexistence of multiple chronic diseases has become increasingly common ^[1, 2]^. The World Health Organization (WHO) defines the coexistence of two or more chronic health problems in the same individual as “multimorbidity” ^[3]^. Globally, the population aged 65 years and older increased from 6% to 9% between 1990 and 2019 and is projected to increase to 16% by 2050^[4]^. In 2023, China’s population aged 60 years and over reached 297 million, accounting for 21.1% of the total population, and is projected to exceed 400 million by 2035, accounting for more than 30% of the total population^[5]^. According to the China Elderly Health Report (2024), the prevalence of chronic diseases such as hypertension, diabetes mellitus, and dyslipidemia has worsened over the past decade, with the prevalence of diabetes mellitus surging by 35%. In the Chinese adult population, the prevalence of multimorbidity has reached 36.1%, and this prevalence continues to show an increasing trend^[6]^. Patients with multimorbidity often require multiple medications, which not only increases the complexity of treatment and care but also leads to a significant rise in adverse events and healthcare costs^[7, 8]^. Of particular concern, rural areas in China have become the hardest hit by multimorbidity due to a severe lack of health resources and a general lack of health awareness among the population^[9]^.

Currently, research on multimorbidity predominantly targets urban communities and older adults aged 60 years and above, while limited attention has been given to multimorbidity patterns in rural populations, particularly younger adults^[6]^. This study focuses on individuals aged 30 years or older in rural areas of North China, aiming to investigate the prevalence of chronic diseases. By collecting real data on multimorbidity in resource-constrained rural areas, we aim to offer a scientific basis and practical recommendations for prevention and management strategies.

## Methods

### Research subjects

This cross-sectional study was conducted between February and December 2023 in rural regions of North China. A whole-cluster stratified random sampling method was employed to select two counties in Shanxi Province, Northern China. Forty villages were subsequently selected from each county using whole-cluster sampling. At least 25 participants were selected from each village, stratified by gender (male/female) and age groups (30–59 years and ≥60 years). Inclusion criteria included being aged ≥30 years and a permanent resident of the selected village. Exclusion criteria comprised pregnant women, individuals unable to provide informed consent, unwilling participants, those unable to communicate effectively, and individuals unable to complete blood and urine sample collection. Ethical approval was obtained from the Ethics Committee of Harbin Medical University (NO. HMUIRB2022005PRE), and all participants provided written informed consent. A total of 2,208 participants were included in the study.

### Research methodology

All study participants were required to complete a questionnaire and undergo a physical examination. The questionnaires were derived from the standardized Assessment of Multimorbidity in Primary Health Care (MAQ-PC) tool^[10]^, covering socio-demographic characteristics, lifestyle habits, and psychosocial factors, while collecting data on variables such as gender, age, marital status, educational level, household size, annual family income, smoking status, alcohol consumption, and physical activity. Alcohol consumption was assessed via the Alcohol Use Disorders Identification Test (AUDIT), where a score of ≥8 indicated hazardous or harmful drinking behavior ^[11]^, Physical activity levels were assessed using the International Physical Activity Questionnaire (IPAQ), classifying participants into low, moderate, or high physical activity categories ^[12]^.

Physical examination included height, weight, waist circumference, and blood pressure. The measurement process adhered to the Chinese population health monitoring anthropometric method standard (WS/T424-2013). For blood pressure measurement, a calibrated Omron HEM-7036 electronic sphygmomanometer was used. Two to three measurements of participants’ sitting upper arm blood pressure were recorded and averaged. Waist circumference ≥90 cm in males and ≥85 cm in females was considered indicative of central obesity^[13]^, while blood pressure ≥130/80 mm Hg was classified as hypertension^[14]^. Body mass index (BMI) was categorized as ≤24 kg/m^2^ for normal weight, 24–28 kg/m^2^ for overweight, and ≥28 kg/m^2^ for obesity.

### Methods of Disease Diagnosis and Types

Disease information in this study was self-reported by the participants and included 19 chronic conditions, such as hypertension (HTN), stroke (CVA), heart disease (HD), arthritis (RA), chronic back pain (CBP), osteoporosis (OP), tuberculosis (TB), chronic lung disease (COPD), chronic digestive disease (CDD), diabetes mellitus (DM), thyroid disease (TD), eye disease (EyeD), ear disease (EarD), epilepsy (EP), anxiety (ANX), depression (MDD), dementia (DEM), chronic kidney disease (CKD), and cancer (CA).

### Sample Size Calculation

The results of a meta-analysis indicated that, over the past 20 years, the prevalence of adult multimorbidity in China was 25.4% (95% CI: 15.1%, 35.7%) ^[15]^ . PASS15.0 software was used to determine the required sample size. A lower prevalence rate of 15.1% was selected, with a two-sided α of 0.05 and a tolerance error of 1.51%. The Simple Asymptotic method was applied to compute a maximum required sample size of 2160 cases. The inclusion of 2208 participants exceeded the calculated sample size requirement.

### Statistical Analysis

Data were independently entered by two individuals and subsequently checked for consistency to create the final database. Statistical analyses were performed using R 4.2.1 software. Count data were described as frequencies, and between-group comparisons were conducted using Pearson’s chi-square test, Fisher’s exact test, or the Kruskal-Wallis test. The multimorbidity network graph was visualized using Gephi 0.10.1 software. Graph density was defined as the ratio of the actual number of edges to the maximum possible number of edges, and the mean degree was calculated as the ratio of the sum of the degrees of all nodes (i.e., number of edges) to the number of nodes (i.e., disease types).In the network graph (**Fig.1**) : Nodes represent different chronic disease types,with their size proportional to the disease prevalence. Nodes corresponding to diseases within the same system are assigned the same color. The thickness of the edges is proportional to the frequency of multimorbidity. Nodes are arranged counterclockwise according to their degree (the number of connected edges).All statistical analyses were deemed significant at P < 0.05.

## Results

### Basic Information

A total of 2208 adults were included in this study. Among them, 1283 (58.11%) were aged 30-59 years, and 1152 (52.17%) were female. Compared to the 30-59 years age group, individuals aged ≥60 years exhibited trends of lower marriage rates, literacy levels, family size, annual household income, smoking rates, high-risk drinking, and high-intensity physical activity, but higher rates of central obesity and systolic blood pressure ≥130 mm Hg. Compared to males, females exhibited higher literacy levels, higher annual household income, but lower rates of smoking, high-risk drinking, and systolic blood pressure ≥130 mm Hg. The proportion of married women and those engaging in high-intensity physical activity were also higher than that of males (**Table 1**).

**Table 1.** Demographic characteristics of adult residents in rural areas of North China (N = 2208)

| Characteristic | Age(n/%) |  | Gender(n/%) |  | Total(n/%) |
| --- | --- | --- | --- | --- | --- |
|  | 30-59 y | ≥60 y | Male | Female |  |
| Total | 1283(58.11) | 925(41.89) | 1056 (47.83) | 1152 (52.17) | 2208(100) |
| Marriage status |  |  |  |  |  |
| Married | 1191 (92.83) | 774 (83.68) * | 917 (86.84) | 1048 (90.97) * | 1965 (88.99) |
| Widowed | 31 (2.42) | 133 (14.38) | 72 (6.82) | 92 (7.99) | 164 (7.43) |
| Others# | 61(4.76) | 18(1.94) | 67(6.34) | 12(1.04) | 79(3.58) |
| Educational level |  |  |  |  |  |
| Primary or less | 491 (38.27) | 523 (56.54) * | 392 (37.12) | 622 (53.99) * | 1014 (45.92) |
| Secondary | 654 (50.97) | 312 (33.73) | 524 (49.62) | 442 (38.37) | 966 (43.75) |
| High school and above | 138 (10.76) | 90(9.73) | 140 (13.26) | 88 (7.64) | 228 (10.33) |
| Number of family members |  |  |  |  |  |
| 1-2 | 296 (23.07) | 501 (54.16) * | 402 (38.07) | 395 (34.29) | 797 (36.10) |
| 3-4 | 614 (47.86) | 139 (15.03) | 357 (33.81) | 396 (34.38) | 753 (34.10) |
| ≥5 | 373 (29.07) | 285 (30.81) | 297 (28.12) | 361 (31.34) | 658 (29.80) |
| Annual family income (Chinese Yuan) |  |  |  |  |  |
| <20000 | 464 (36.17) | 633 (68.43) * | 528 (50.00) | 569 (49.39) * | 1097 (49.68) |
| 20000-50000 | 608 (47.39) | 221 (23.89) | 376 (35.61) | 453 (39.32) | 829 (37.55) |
| ≥50000 | 211 (16.45) | 71 (7.68) | 152 (14.39) | 130 (11.28) | 282 (12.77) |
| Smoke status |  |  |  |  |  |
| Never smoked | 828 (64.54) | 621 (67.14) * | 311 (29.45) | 1138 (98.78) * | 1449 (65.62) |
| Used to smoke | 40 (3.12) | 68 (7.35) | 106 (10.04) | 2 (0.17) | 108 (4.89) |
| Now smoking | 415 (32.35) | 236 (25.51) | 639 (60.51) | 12 (1.04) | 651 (29.48) |
| Drinking alcohol (using AUDIT) |  |  |  |  |  |
| Low-risk drinking | 936 (72.95) | 755 (81.62) * | 569 (53.88) | 1122 (97.40) * | 1691 (76.59) |
| High-risk drinking | 347 (27.05) | 170 (18.38) | 487 (46.12) | 30 (2.60) | 517 (23.41) |
| Physical activity (using IPAQ) |  |  |  |  |  |
| Low intensity | 128 (9.98) | 120 (12.97) * | 118 (11.17) | 130 (11.28) * | 248 (11.23) |
| Medium intensity | 158 (12.31) | 157 (16.97) | 182 (17.23) | 133 (11.55) | 315 (14.27) |
| High intensity | 997 (77.71) | 648 (70.05) | 756 (71.59) | 889 (77.17) | 1645 (74.50) |
| BMI (kg/m<sup>2</sup>) |  |  |  |  |  |
| ≤24 | 433 (33.75) | 383 (41.41) * | 406 (38.45) | 410 (35.59) | 816 (36.96) |
| 24-28 | 547 (42.63) | 364 (39.35) | 427 (40.44) | 484 (42.01) | 911 (41.26) |
| ≥28 | 303 (23.62) | 178 (19.24) | 223 (21.12) | 258 (22.40) | 481 (21.78) |
| Waist circumference |  |  |  |  |  |
| Normal | 479 (37.33) | 287 (31.03) * | 386 (36.55) | 380 (32.99) | 766 (34.69) |
| Central obesity | 804 (62.67) | 638 (68.97) | 670 (63.45) | 772 (67.01) | 1442 (65.31) |
| SBP (mm Hg) |  |  |  |  |  |
| <130 | 660 (51.44) | 322 (34.81) * | 445 (42.14) | 537 (46.61) * | 982 (44.47) |
| ≥130 | 623 (48.56) | 603 (65.19) | 611 (57.86) | 615 (53.39) | 1226 (55.53) |
| DBP (mm Hg) |  |  |  |  |  |
| <80 | 436 (33.98) | 398 (43.03) * | 340 (32.20) | 494 (42.88) * | 834 (37.77) |
| ≥80 | 847 (66.02) | 527 (56.97) | 716 (67.80) | 658 (57.12) | 1374 (62.23) |

\*Statistically significant at 5 level.
#Others denotes divorced/separated/unmarried.

### Prevalence of Chronic Diseases

The prevalence of chronic diseases was 66.53%. The most common conditions were hypertension (HTN, 43.21%), coronary artery disease (CDD, 11.82%), cerebrovascular accident (CVA, 10.19%), rheumatoid arthritis (RA, 10.10%), and diabetes mellitus (DM, 9.78%). The prevalence was significantly higher among individuals aged ≥60 years compared to those aged 30-59 years (82.27% vs. 55.18%, P<0.05). There was no significant difference in prevalence between males and females (64.96% vs. 67.97%, P>0.05) (**Table 2**).

**Table 2.** The prevalence of 19 chronic diseases in 2208 adults by sex and age.

| Chronic diseases | Age(n/%) |  | Gender(n/%) |  | Total(n/%) |
| --- | --- | --- | --- | --- | --- |
|  | 30-59y(n=1283) | ≥60y(n=925) | Male(n=1056) | Female(n=1152) |  |
| HTN | 411(32.03) | 543(58.70) * | 448(42.42) | 506(43.92) | 954(43.21) |
| CDD | 129(10.05) | 132(14.27) * | 108(10.23) | 153(13.28) * | 261(11.82) |
| CVA | 70(5.46) | 155(16.76) * | 140(13.26) | 85(7.38) * | 225(10.19) |
| RA | 98(7.64) | 125(13.51) * | 60(5.68) | 163(14.15) * | 223(10.10) |
| DM | 80(6.24) | 136(14.70) * | 88(8.33) | 128(11.11) * | 216(9.78) |
| HD | 62(4.83) | 138(14.92) * | 104(9.85) | 96(8.33) | 200(9.06) |
| CBP | 72(5.61) | 68(7.35) | 61(5.78) | 79(6.86) | 140(6.34) |
| EyeD | 17(1.33) | 76(8.22) * | 33(3.12) | 60(5.21) * | 93(4.21) |
| COPD | 35(2.73) | 50(5.41) * | 49(4.64) | 36(3.12) | 85(3.85) |
| EarD | 24(1.87) | 38(4.11) * | 39(3.69) | 23(2.00) * | 62(2.81) |
| CA | 28(2.18) | 33(3.57) | 19(1.80) | 42(3.65) * | 61(2.76) |
| TD | 34(2.65) | 26(2.81) | 13(1.23) | 47(4.08) * | 60(2.72) |
| OP | 25(1.95) | 32(3.46) * | 20(1.89) | 37(3.21) | 57(2.58) |
| CKD | 18(1.40) | 14(1.51) | 18(1.70) | 14(1.22) | 32(1.45) |
| TB | 12(0.94) | 9(0.97) | 6(0.57) | 15(1.30) | 21(0.95) |
| EP | 8(0.62) | 2(0.22) | 7(0.66) | 3(0.26) | 10(0.45) |
| ANX | 6(0.47) | 2(0.22) | 2(0.19) | 6(0.52) | 8(0.36) |
| DEM | 1(0.08) | 6(0.65) * | 5(0.47) | 2(0.17) | 7(0.32) |
| MDD | 6(0.47) | 1(0.11) | 2(0.19) | 5(0.43) | 7(0.32) |
| Total | 708(55.18) | 761(82.27) * | 686(64.96) | 783(67.97) | 1469(66.53) |

\*Statistically significant at 5% level.

### Status of multimorbidity

The prevalence of local multimorbidity (defined as having two or more chronic diseases simultaneously) was 32.47%. Multimorbidity was more common in people aged ≥60 years than in those aged 30-59 years (47.68% vs. 21.51%, P<0.05). No statistically significant difference in multimorbidity prevalence was found between males and females (30.87% vs. 33.94%, P>0.05).

Among the 717 cases of multimorbidity, more than 55% had two concurrent diseases, approximately one-quarter had three diseases, and nearly one-fifth had four or more diseases.

The number of chronic conditions reached up to 10 in some cases. Hypertension (HTN) was the most common comorbidity, affecting 78.52% of individuals (**Table 3**).

**Table 3.**
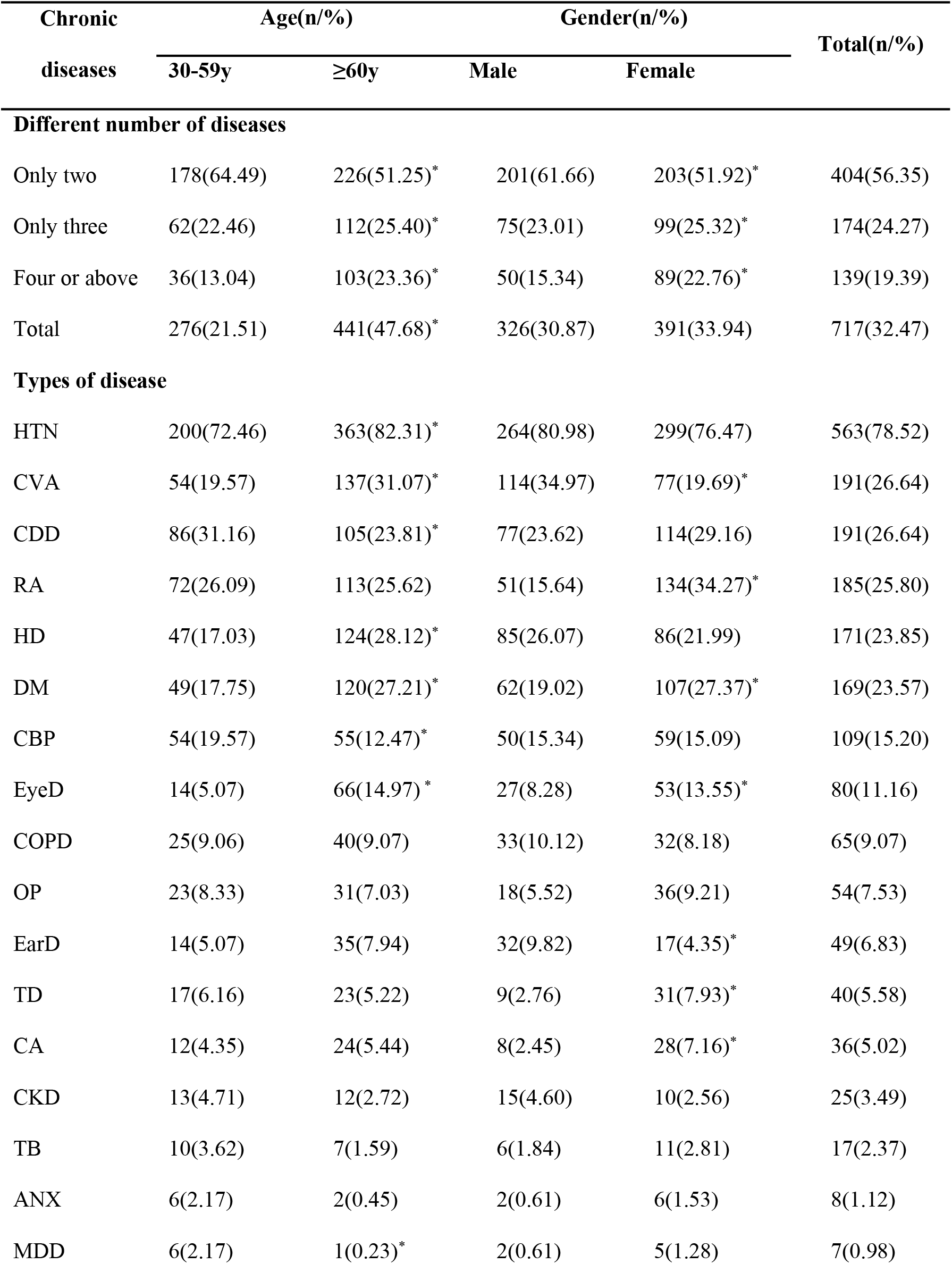
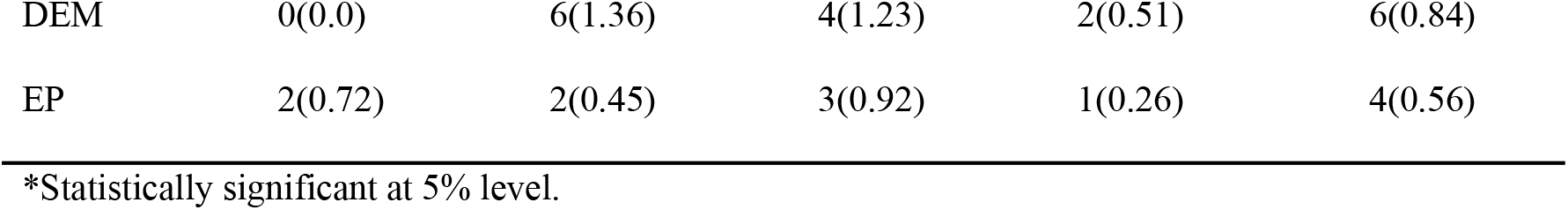
The distribution of 19 chronic conditions among 717 cases with multimorbidity, stratified by age and gender.

### Analysis of Chronic Disease Multimorbidity Patterns

The top three dyad disease patterns in this study were HTN&CVA (7.47%), HTN&HD (6.25%), and HTN&DM (6.11%). HTN&HD&CVA (1.77%) was the most common triad disease pattern. In the 30-59-year-old group, the top three dyad disease patterns were HTN&CDD (3.82%), HTN&CVA (3.74%), and HTN&RA (3.43%). Among those aged ≥60 years, HTN&CVA (12.65%), HTN&HD (11.46%), and HTN&DM (10.38%) were the most common patterns, as shown in **Table 4**. For males, the most common dyad disease patterns were HTN&CVA (9.47%), HTN&HD (6.53%), and HTN&CDD (4.73%). For females, HTN&RA (8.16%), HTN&DM (7.64%), and HTN&CDD (6.77%) were the most prevalent, as shown in **Table 5**.

**Table 4.**
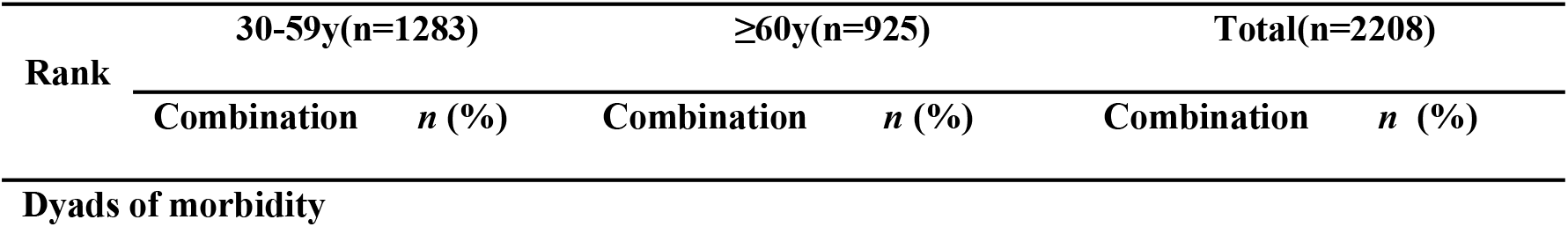
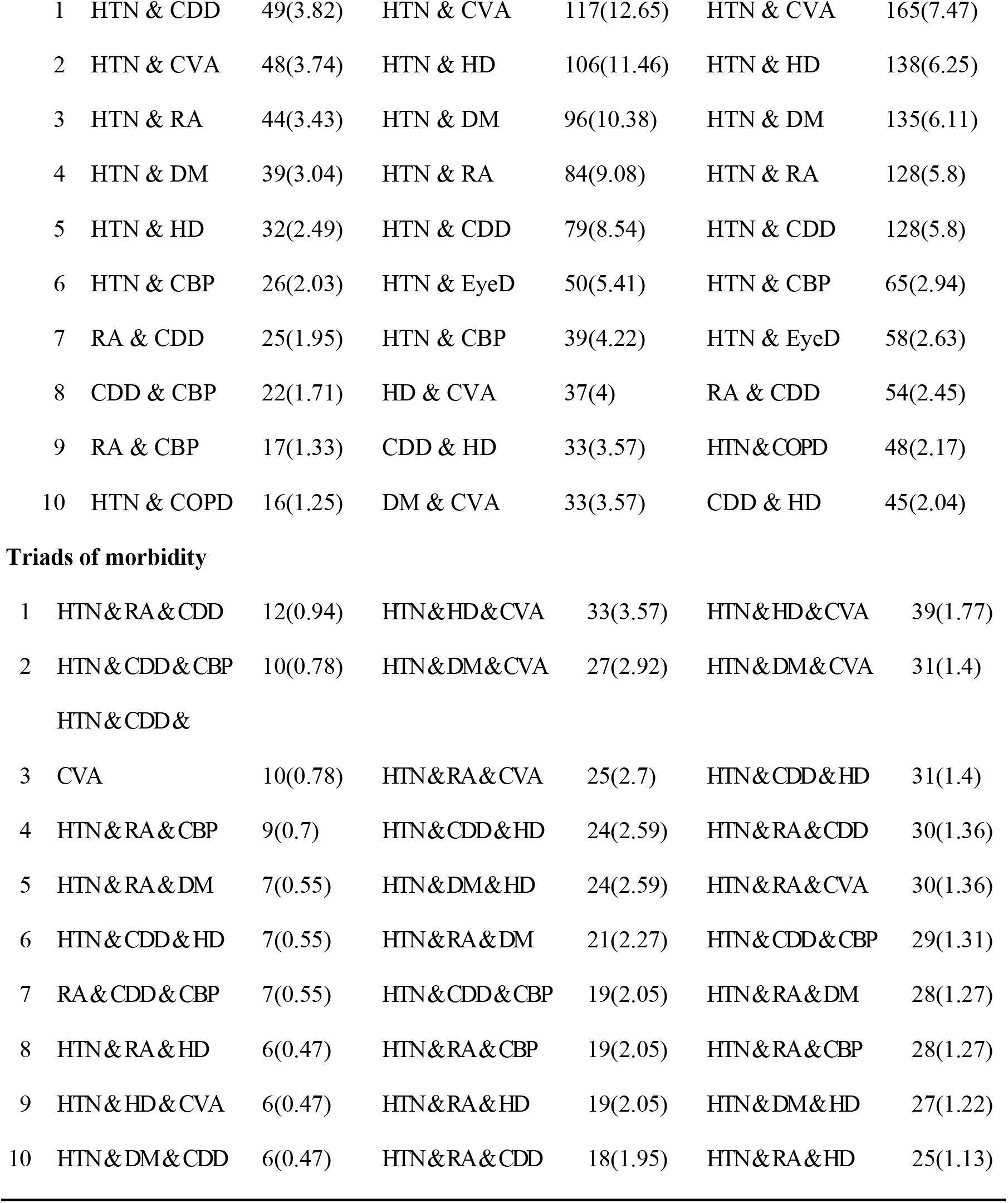
Top ten frequent unique combination clusters with multimorbidity, stratified by age.

|  |  |  |  |  |  |  |
| --- | --- | --- | --- | --- | --- | --- |
| 1 | HTN & CDD | 49(3.82) | HTN & CVA | 117(12.65) | HTN & CVA | 165(7.47) |
| 2 | HTN & CVA | 48(3.74) | HTN & HD | 106(11.46) | HTN & HD | 138(6.25) |
| 3 | HTN & RA | 44(3.43) | HTN & DM | 96(10.38) | HTN & DM | 135(6.11) |
| 4 | HTN & DM | 39(3.04) | HTN & RA | 84(9.08) | HTN & RA | 128(5.8) |
| 5 | HTN & HD | 32(2.49) | HTN & CDD | 79(8.54) | HTN & CDD | 128(5.8) |
| 6 | HTN & CBP | 26(2.03) | HTN & EyeD | 50(5.41) | HTN & CBP | 65(2.94) |
| 7 | RA & CDD | 25(1.95) | HTN & CBP | 39(4.22) | HTN & EyeD | 58(2.63) |
| 8 | CDD & CBP | 22(1.71) | HD & CVA | 37(4) | RA & CDD | 54(2.45) |
| 9 | RA & CBP | 17(1.33) | CDD & HD | 33(3.57) | HTN&COPD | 48(2.17) |
| 10 | HTN & COPD | 16(1.25) | DM & CVA | 33(3.57) | CDD & HD | 45(2.04) |
| Triads of morbidity |  |  |  |  |  |  |
| 1 | HTN&RA&CDD | 12(0.94) | HTN&HD&CVA | 33(3.57) | HTN&HD&CVA | 39(1.77) |
| 2 | HTN&CDD&CBP | 10(0.78) | HTN&DM&CVA | 27(2.92) | HTN&DM&CVA | 31(1.4) |
|  | HTN&CDD& |  |  |  |  |  |
| 3 | CVA | 10(0.78) | HTN&RA&CVA | 25(2.7) | HTN&CDD&HD | 31(1.4) |
| 4 | HTN&RA&CBP | 9(0.7) | HTN&CDD&HD | 24(2.59) | HTN&RA&CDD | 30(1.36) |
| 5 | HTN&RA&DM | 7(0.55) | HTN&DM&HD | 24(2.59) | HTN&RA&CVA | 30(1.36) |
| 6 | HTN&CDD&HD | 7(0.55) | HTN&RA&DM | 21(2.27) | HTN&CDD&CBP | 29(1.31) |
| 7 | RA&CDD&CBP | 7(0.55) | HTN&CDD&CBP | 19(2.05) | HTN&RA&DM | 28(1.27) |
| 8 | HTN&RA&HD | 6(0.47) | HTN&RA&CBP | 19(2.05) | HTN&RA&CBP | 28(1.27) |
| 9 | HTN&HD&CVA | 6(0.47) | HTN&RA&HD | 19(2.05) | HTN&DM&HD | 27(1.22) |
| 10 | HTN&DM&CDD | 6(0.47) | HTN&RA&CDD | 18(1.95) | HTN&RA&HD | 25(1.13) |

**Table 5.**
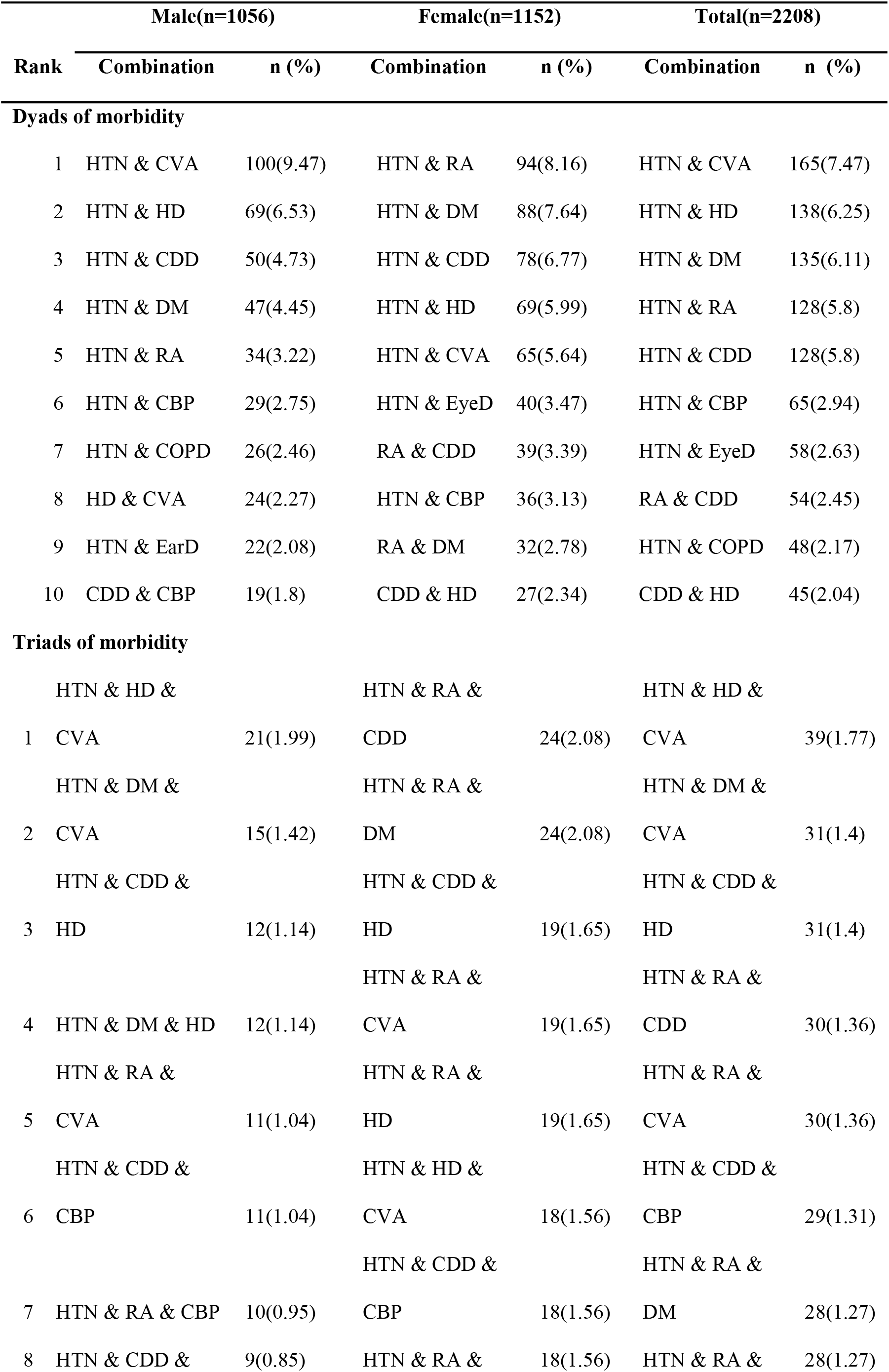
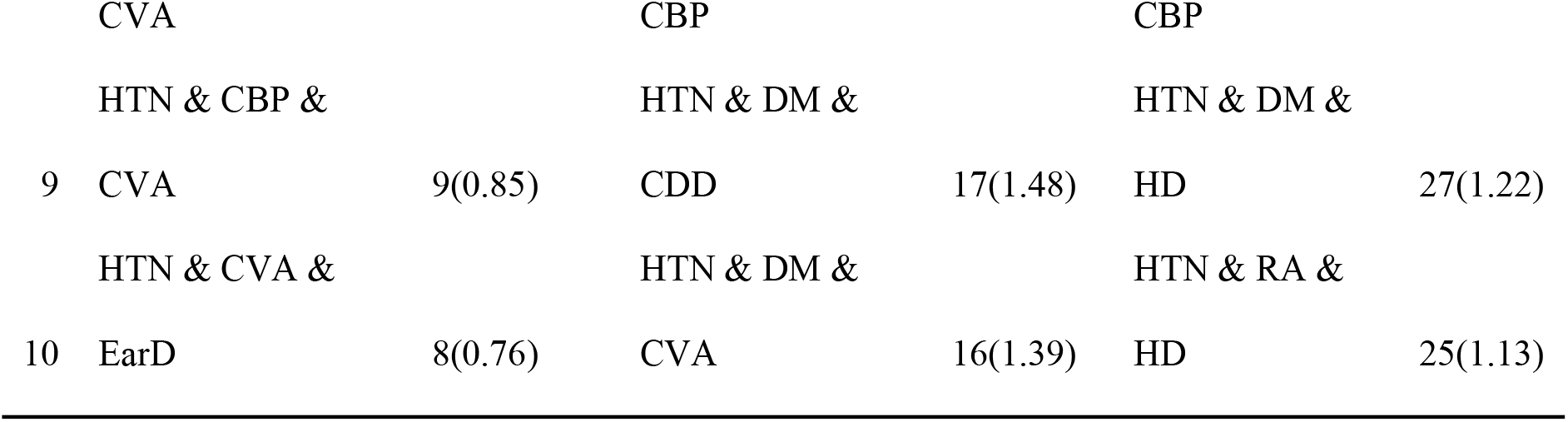
Top ten frequent unique combination clusters with multimorbidity, stratified by gender.

In the network graph of the total population (**Fig. 1a**), 137 contiguous edges were observed, with a graph density of 0.80 and a mean degree of 14.42. In the network graph of the 30-59-year-old population (**Fig. 1b**), 103 contiguous edges were observed, with a graph density of 0.60 and a mean degree of 10.84. In the network graph for those aged ≥60 years (**Fig. 1c**), 125 edges were observed, with a graph density of 0.73 and a mean degree of 13.16. The ≥60-year-old group exhibited greater graph density and thicker edges. In the male population network graph (**Fig. 1d**), 108 contiguous edges were observed, with a graph density of 0.63 and a mean degree of 11.37. In the female population network graph (**Fig. 1e**), 122 edges were observed, with a higher graph density of 0.71 and a mean degree of 12.84. Females exhibited greater graph density and thicker edges compared to males.

**Fig. 1.**
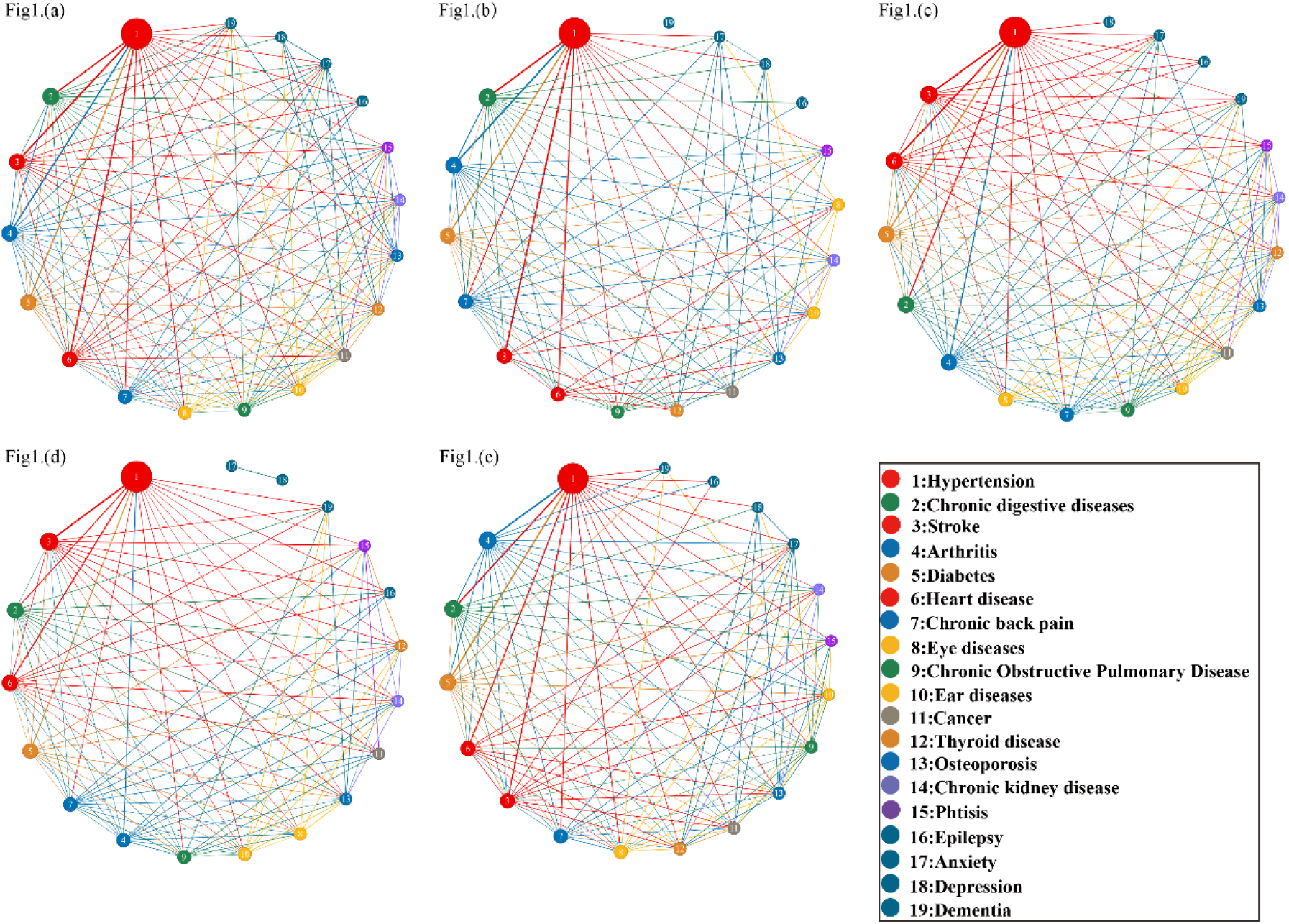
presents a network model diagram illustrating multimorbidity patterns across different. age groups and genders: (a) overall population; (b) individuals aged 30-59 years; (c) individuals aged 60 years and above; (d) male population; and (c) female population.

## Discussion

In 2023, the prevalence of chronic diseases among individuals aged 30 years and older in rural northern China was 66.53%, and the multimorbidity rate was 32.47%. Hypertension (HTN) was the most common chronic disease, with a prevalence of 43.21%. The prevalence and multimorbidity rates were significantly higher among individuals aged 60 years and older compared to those aged 30-59 years, with no significant differences observed between genders. The combination of hypertension with other chronic diseases was common, particularly the combinations of HTN & CVA, HTN & HD, and HTN & DM. These multimorbidity were prevalent across both genders and various age groups.

The prevalence of chronic diseases in this study (66.53%) was notably higher than that reported in the CHARLS study in China in 2018 (50.00%)^[16]^, in rural areas of Southwest China in 2021 (50.01%)^[17]^, and in rural Iran in 2019 (49.6%)^[18]^. Several factors may explain the increase in chronic disease prevalence in rural China in recent years. First, improved economic conditions and more refined diets, combined with limited health knowledge about scientific diets, have contributed to an imbalance between energy intake and expenditure, leading to increased rates of overweight and obesity ^[19, 20]^. Second, mechanization has replaced traditional farming methods, and the widespread use of electronic devices has led to a significant reduction in physical activity and exercise^[21]^. Additionally, advancements in China’s healthcare system and policies that increase medical insurance coverage for chronic diseases have extended life expectancy^[22]^.Finally, the inclusion of a broader range of chronic diseases in this study may also contribute to the higher prevalence rates. The prevalence of multimorbidity in rural China reported in this study was higher than that of the CHARLS study in China (31.53%)^[23]^but lower than global (37.2%) and Asian (35%) averages^[24]^. The prevalence of chronic diseases among adults aged 30-60 years in this study was 55.18%, indicating that this age group will likely be a major contributor to future multimorbidity growth in rural China. This finding suggests that multimorbidity is increasing rapidly in rural China and will likely contribute to a significant disease burden in these areas.

Various factors, including geographic environment, lifestyle, medical resource distribution, genetic background, and socioeconomic conditions^[25]^, influence the focus of multimorbidity studies across different countries and regions. For instance, in the United States, hyperlipidemia, hypertension, and diabetes mellitus are the main multimorbid conditions, underscoring the predominance of metabolic disease multimorbidity^[26]^. In France, alongside metabolic diseases, mental health conditions are also prevalent^[27]^. This study, however, included a broader range of chronic diseases, such as circulatory and metabolic disorders (e.g., hypertension, stroke, diabetes), as well as digestive, respiratory, musculoskeletal, and mental health conditions.

Hypertension was the most prevalent condition among individuals with multimorbidity in rural China, consistent with previous studies^[28]^. This may be associated with dietary habits, such as high salt intake, a carbohydrate-based diet, and insufficient consumption of fresh fruits and vegetables in rural northern China^[29]^. Hypertension, stroke, diabetes mellitus, and other cardiovascular and metabolic diseases were commonly observed as multimorbid conditions. Hypertension, in particular, was strongly linked to the onset of these diseases^[30]^, underscoring its critical role in chronic disease management^[31]^.

In this study, chronic disease multimorbidity was stratified by age and gender, with the most common dyad disease patterns among individuals aged 30–59 years being the combination of HTN with CDD, CVA, and RA. The high prevalence of CDD in this age group is closely related to irregular dietary habits, lack of exercise, high stress, and other factors. Especially since this region has a high prevalence of digestive tumors, such as esophageal cancer^[32]^, a possible genetic effect may be at play. The combination of CVA is consistent with its earlier onset, likely due to the high prevalence of risk factors such as hypertension, hyperlipidemia, obesity, and smoking in younger populations^[33]^. The combination of RA may be linked to the cold climate in northern rural areas, prolonged periods of insufficient sunlight, and a mono-diet leading to inadequate intake of calcium and other micronutrients^[34]^. In individuals over 60 years of age, the combination of HTN with CVA, HD, and DM is predominantly associated with the cumulative onset of age-related diseases, including circulatory and metabolic disorders^[35, 36]^. Among males, common dyad combinations include HTN with CVA, HD, and CDD. Smoking and excessive alcohol consumption have been identified as key behavioral risk factors for cardiovascular disease in men^[37]^. In contrast, the most frequent combinations among females are HTN with RA, DM, and CDD. Studies indicate that females are at higher risk of chronic diseases such as RA and DM compared to males^[38, 39]^, potentially due to the stronger influence of altered estrogen levels and psychosocial risk factors^[40]^.

The study provided comprehensive coverage of disease types and ensured the completeness of the data and the reliability of the findings. However, the representativeness of the data may be limited because the study was conducted exclusively in rural areas of a single province.

Nevertheless, the findings remain informative due to the similarities in demographics, age distribution, lifestyle habits, and the prevalence of chronic diseases between rural areas of Shanxi Province and other northern provinces in China. In addition, the study analyzed the effects of gender and age group on multimorbidity, but future research should consider the influence of other demographic characteristics, lifestyle factors, and physical examination variables on the combination of multimorbidity.

## Conclusions

This study reveals the severity of chronic diseases and multimorbidity among adults in rural areas of northern China, with hypertension identified as the most prevalent chronic condition contributing to multimorbidity. Various demographic groups exhibit distinct health characteristics, necessitating stratified, precise, and standardized management approaches. For instance, individuals aged 30-59 years should be managed for hypertension and chronic digestive diseases; those aged 60 years or older should receive management for hypertension alongside conditions like stroke, heart disease, and diabetes mellitus. Males should be monitored for hypertension and chronic digestive diseases, while females require management for hypertension and arthritis.

## Data Availability

Data are available from the Corresponding authors

## List of Abbreviations

HTN: hypertension
CVA: stroke
HD: heart disease
RA: arthritis
CBP: chronic back pain
OP: osteoporosis
TB: tuberculosis
COPD: chronic lung disease
CDD: chronic digestive disease
DM: diabetes mellitus
TD: thyroid disease
EyeD: eye disease
EarD: ear disease
EP: epilepsy
ANX: anxiety
MDD: depression
DEM: dementia
CKD: chronic kidney disease
CA: and cancer
IPAQ: the International Physical Activity Questionnaire
AUDIT: Alcohol Use Disorders Identification Test

## Declarations

### Ethics approval and consent to participate

The study was approved by the Ethics Committee of Harbin Medical University (NO. HMUIRB2022005PRE) before its commencement and conducted in accordance with the guidelines outlined in the Declaration of Helsinki. Informed consent was obtained from all participants before their involvement in the study.

### Consent for publication

Not applicable.

### Availability of data and materials

The data from Shanxi, China, needs to be obtained by sending a data request application to the corresponding author’s email.

### Competing interests

The authors declare no competing interests.

### Funding

Chengzhi Medical College Doctoral Startup Fund, Grant/Award Number: 2024BS14.

### Authors’ Information

Affiliations

School of Public Health, Shanxi Medical University, Taiyuan 030001, China. ShuaiTang, Yanxing Li, Mei li Niu, Zijing Qi & Zhifang Li

Department of Prevention and Health Care, Affiliated Heping hospital of Changzhi medical college, Changzhi, 046000, China.

Tianyou Hao

Department of Public Health and Prevention, Changzhi Medical College, Changzhi 046000, China.

Hongmei Yang, Xiangxian Feng & Zhifang Li

The George Institute for Global Health, University of New South Wales, Australia. Maoyi Tian

School of Medical Board, Shanxi Datong University, Datong 037009, China. Maoyi Tian & Xinyi Zhang

### Contributions

S.T., L.Y. and Z.L; methodology, M.T.,X.F. and Z.L; software, S.T., L.Y. and Z.Q; validation, H.Y., Y.L., T.H., M.N., Z.Q and S.T.; investigation, Z.X., S.T., T.H., H.Y. and M.N.; resources, T.H., H.Y.; data curation, S.T. and Y.L.; writing—original draft preparation, S.T. and Y.L.; writing—review and editing, S.T., Y.L., Y.H.,X.Z, M.T., X.F. and Z.L.; visualization, T.H., Y.H. and F.X.; supervision, M.T., X.F. and Z.L.; project administration, X.F. and Z.L.; funding acquisition, Z.L. All authors have read and agreed to the published version of the manuscript.

### Corresponding authors

Correspondence to Zhifang Li.

## Notes

### Competing Interest Statement

The authors have declared no competing interest.

### Funding Statement

The author(s) received no specific funding for this work.

### Author Declarations

Ethical approval was obtained from the Ethics Committee of Harbin Medical University (NO. HMUIRB2022005PRE), and all participants provided written informed consent.

